# Efficacy and Safety of an Inactivated Whole-Virion SARS-CoV-2 Vaccine (CoronaVac) in Brazilian Healthcare Professionals: The PROFISCOV Trial

**DOI:** 10.1101/2024.04.22.24306197

**Authors:** José Moreira, Elizabeth G. Patiño, Patricia Emilia Braga, Pedro Pacheco, Caroline Curimbaba, Christopher Gast, Ricardo Palácios, Mauro Teixeira, Fabiano Ramos, Gustavo Romero, Fabio Leal, Luiz Junior, Luiz Camargo, Francisco Aoki, Eduardo Coelho, André Siqueira, Sonia Raboni, Danise Oliveira, Paulo Tarso, Cor Fontes, Ana Lyrio, Mauricio Nogueira, Fernanda Boulos, Esper Kallas, PROFISCOV study group

**Affiliations:** Clinical Trials and Pharmacovigilance Center, Instituto Butantan, São Paulo, Brazil; PATH, Center for Innovation and Access, Seattle, WA, USA; GSK, Siena, Italy; Faculdade de Medicina, Universidade Federal de Minas Gerais (UFMG), Belo Horizonte, Brazil; Hospital São Lucas da PUC, Porto Alegre, Brazil; Universidade de Brasileira, Brasília, Brazil; Universidade de São Caetano do Sul, São Paulo, Brazil; Instituto de Infectologia Emilio Ribas, São Paulo, Brazil; Hospital Albert Einstein, São Paulo, Brazil; Universidade de Campinas, Campinas, Brazil; Hospital das Clínicas da Faculdade de Medicina de Ribeirão Preto da Universidade de São Paulo, Ribeirão Preto, Brazil; Instituto Nacional de Infectologia Evandro Chagas, Fundação Oswaldo Cruz, Rio de Janeiro, Brazil; Complexo Hospital das Clínicas da Universidade Federal do Paraná (EBSERH), Curitiba, Brazil; Centro de Pesquisa Clínica do Hospital Escola da Universidade Federal de Pelotas, Pelotas, Brazil; Hospital de Amor Nossa Senhora (HANS), Fundação PIO XII, Barretos, Brazil; Hospital Universitário Júlio Muller, Cuiabá, Brazil; Universidade Federal de Mato Grosso do Sul (FAMED), Hospital Universitário Maria Aparecida Pedrossian, Campo Grande, Brazil; Faculdade de Medicina de São José do Rio Preto (FAMERP), São José do Rio Preto, Brazil

**Author notes:** Corresponding author: José Moreira, M.D, Ph.D., Clinical Developmental Medical Director, Clinical Trials and Pharmacovigilance Center, Instituto Butantan. Av. Vital Brasil 1500. São Paulo, Brazil, 05503-900. Members of the PROFISCOV study group are listed in the Supplementary Appendix. Part of this work was presented at the 2023 American Society of Tropical Medicine & Hygienés Annual Meeting in Chicago, Illinois, USA.

## Abstract

**Background:** CoronaVac, an inactivated COVID-19 vaccine, underwent evaluation for its efficacy and safety during the PROFISCOV study conducted in Brazil.

**Methods:** Between July 21, 2020, and July 29, 2021, 13,166 participants provided informed consent, with 12,688 randomized for the trial. Participants were allocated between vaccine and placebo arms (1:1) and monitored for symptomatic COVID-19 cases, severity of disease, and adverse reactions after two doses given 14 days apart.

**Findings:** The primary efficacy analysis revealed a vaccine efficacy of 50□39% (95% confidence interval [CI], 35·26% to 61·98%; *p=*0·0049) in preventing symptomatic COVID-19, leading to the issuance of Emergency Use Authorization for CoronaVac in January 2021. Upon completion of follow-up, vaccine efficacy was 44□58% [95% CI, 34·89% to 52·83%; *p=* 0·0023] in preventing COVID-19 and 82□14% (95% CI, 64·93% to 90·90%; *p<*0·0001) in preventing severe COVID-19. Safety data indicated that adverse reactions were more frequent in the vaccine arm, primarily mild to moderate, with pain at the injection site and headache being the most common.

**Interpretation:** CoronaVac demonstrated moderate efficacy in preventing symptomatic COVID-19 and high efficacy against severe disease. While reactions were slightly more common in the vaccine group, they were generally mild and manageable.

**Funding:** Fundação Butantan, Instituto Butantan, and São Paulo Research Foundation (FAPESP; Grants 2020/10127-1 and 2020/06409-1).

**Research in Context:** *Evidence before this study:* At the time of the studýs design in 2020, the world was grappling with the COVID- 19 pandemic, with no licensed vaccine available. A global race to develop a safe and effective vaccine was underway, leading to the exploration of several vaccine candidates based on various technologies and mechanisms of action. Among these candidates was CoronaVac, an inactivated vaccine developed by Sinovac Life Sciences. PubMed was searched for pre-clinical and clinical trials using terms “COVID-19”, “SARS-CoV-2”, “Vaccine”, “Vaccine Efficacy”, without language or data restrictions. Additionally, information on clinical trials was sought from the ClinicialTrials.gov database and regulatory agencies. The focus was on late-stage clinical trials evaluating the safety, immunogenicity, and efficacy of CoronaVac. Positive safety and immunogenicity results from phase I/II clinical trials in younger and older adults, coupled with expanding pandemic, motivated the design and implementation of this phase III trial in healthcare professionals directly caring for or likely to be in close contact with COVID-19 patients in Brazil. No previous phase III study focusing on the efficacy and safety of CoronaVac in this high-risk population was identified.

*Added value of this study:* Between July 21, 2020, and July 29, 2021, 12,688 participants were randomized to receive either CoronaVac or placebo. We evaluated symptomatic COVID-19 cases, disease severity, and adverse reactions after two doses given 14 days apart. We found that CoronaVac met the predefined efficacy criteria, providing a moderate efficacy against symptomatic COVID-19 of 50□39% (95% CI: 35·26-61·98) in the primary analysis. Notably, CoronaVac demonstrated high effective against severe disease, with a vaccine efficacy of 82□14% (64·93-90·90) in the final analysis. Regarding safety, CoronaVac was shown to be safe, with most reactions being mild and manageable, albeit more commonly reported in the CoronaVac group. The inclusion of a high-risk study population comprising healthcare workers directly involved in the care of COVID-19 patients in Brazil is a key differentiator of our trial, as other studies of CoronaVac in China, Indonesia, Chile and Turkey at that time were not restricted to healthcare workers.

*Implications of all the available evidence.:* The primary efficacy analysis data from this study supported the Emergency Use Authorization issued for CoronaVac in Brazil in January 2021. Subsequently, a national vaccination campaign was initiated, with CoronaVac being the first vaccine to be incorporated in the COVID-19 vaccination program in Brazil. Since then, more than 100 million doses of CoronaVac have been administered in Brazil through the National Health System. The efficacy and safety of two doses of CoronaVac were demonstrated in the final analysis of the study. CoronaVac’s ability to prevent severe disease is a crucial attribute that has had a positive impact on pandemic control and public health. It represents a promising option for COVID-19 vaccination, especially in low- or middle-income countries, given its moderate efficacy against symptomatic disease and favorable safety profile, in addition to its lower cost and ease of manufacturing compared to other vaccines available early in the pandemic. The impact on the immunogenicity and safety profile of XBB-updated versions of the vaccine used as a booster vaccination needs to be investigated in future studies.

## INTRODUCTION

The global imperative to contain the Coronavirus 2019 (COVID-19) pandemic, caused by transmission of severe acute respiratory syndrome coronavirus 2 (SARS-CoV-2), incited unprecedented endeavors to formulate, test, and distribute effective vaccines.^1^ Amid the vast array of SARS-CoV-2 vaccine platforms, encompassing purified inactivated virus, recombinant subunits, adenovirus-based vectors, and DNA- or RNA-based formulations, one of the most widely deployed vaccine, particularly early in the acute phase of pandemic in low-and middle-income countries was CoronaVac, an inactivated whole-virion vaccine developed by Sinovac Life Sciences.^2^ CoronaVac demonstrated promising results in preclinical studies, and demonstrated encouraging safety and immunogenicity from phase 1 to 3 clinical trials involving adults.^3–8^ While phase 3 trials took place in countries like Chile, Indonesia, China, and Turkey, none of these trials were exclusively confined to a cohort of healthcare workers (HCW), and conclusive findings among this study population remain unpublished. Data indicate that HCWs early on the COVID-19 pandemic were initially at higher risk of acquiring infection than the general population, and there was also a risk of onward transmission to patients who were at higher risk of serios COVID-19 outcomes through their contact with these worksers.^9^

During the initial 2-years of the pandemic, Brazil grappled with one of the most substantial burdens of COVID-19.^10^ Nonetheless, it has been estimated that, within the first year of the vaccination campaigńs initiation in January 2021, the lives of at least 303,129 adults in the country were preserved.^11^ The present phase 3 trial was designed to assess the safety and efficacy of CoronaVac among HCW engaged in direct care for COVID-19 patients in Brazil. Our hypothesis was that CoronaVac could mitigate both the occurrence and severity of COVID-19 among Brazilian HCWs in comparison with those who received a placebo.

Within this context, we present the outcomes of the primary analysis of vaccine efficacy, conducted with a data cutoff of December 16, 2020 – a pivotal developmental that culminated with the issuance of Emergency Use Authorization (EUA) for CoronaVac by the Brazilian Health Regulatory Agency (ANVISA) in January 2021. Furthermore, we furnish the updated results encompassing efficacy and safety from the final analysis, incorporating data until July 29, 2021. These results represent a comprehensive panorama of the efficacy and safety profile of CoronaVac within a high-risk population, serving as a vital compass for shaping public health policy and refining vaccination strategies.

## METHODS

### Study design

The PROFISCOV trial (NCT0445659) was a double-blind, randomized, placebo-controlled phase 3 trial.^12^ This trial was designed and coordinated by Instituto Butantan in São Paulo, Brazil, in collaboration with other participating institutions and the developer of CoronaVac (Sinovac Life Sciences, Beijing, China). A complete list of the participating institutions and investigators is detailed in the Supplementary Material. Funding for the trial was sourced from Instituto Butantan, a public institution affiliated with the São Paulo State Secretariat of Health. The protocol and its three amendments were reviewed and approved by the local Ethics Review Board, the Brazilian National Research Ethics Council (CONEP), and ANVISA. An independent Data and Safety Monitoring Board (DSMB) monitored participant safety and assessed results of the interim analyses. An external Clinical Endpoint Adjudication Committee (CEAC), comprising medical experts in the field of vaccine development, conducted blinded review of each potential primary endpoint case, including status as severe or not severe, and was responsible for adjudication of suspected COVID-19 cases; only confirmed cases contributed to the efficacy analysis.

### Participants

Eligible participants were HCW aged ≥ 18 years directly engaged in the care of patients with possible or confirmed COVID-19. Initially, individuals with a history of SARS-CoV-2 infection were excluded, but this criterion was revised in the second protocol amendment, which allowed individuals with a history of sero-exposure to SARS-CoV-2 to enroll. Female participants underwent a pregnancy test at baseline and were to adhere to effective contraception for 3-months post-vaccination. Notably, individuals with uncontrolled serious diseases, bleeding disorders, or asplenia were excluded, as were those utilizing immunosuppressive, antineoplastic treatments, or recent blood products. Further exclusions comprised participants with behavioral, cognitive, or psychiatric illness, alcohol or substance abuse, recent vaccination, prior COVID-19 vaccination, or recent fever. The detailed eligibility criteria are provided in the Protocol (see Supplementary Material). Written informed consent was secured from all participants prior to study enrollment.

### Randomization and masking

Participants were randomly assigned 1:1 to receive CoronaVac or placebo, employing permuted blocks and stratifying by age (18-59 *vs*. ≥ 60 years). A uniform randomization list was accessible to all sites via an interactive web-based response system provided by Cenduit (Durham, NC). The masking protocol was meticulously implemented, involving participants, trial personnel (including physicians, nurses, monitors, laboratory technicians, and the data management team), and unmasked pharmacists providing syringes to masked research personnel responsible for administration and observation. The placebo was manufactured under Good Manufacture Practices (GMP) and resembled the active product. An EUA for CoronaVac was granted by ANVISA on January 17, 2021 based on the primary efficacy analysis, leading to protocol amendments and the unblinding of participants, in compliance with ethical standards.

### Procedures

CoronaVac was formulated from the CN02 strain of SARS-CoV-2 propagated in African green monkey kidney (Vero) cells. The virus was inactivated with β- propiolactone and affixed to aluminum hydroxide.^3^ Placebo comprised aluminum hydroxide diluent devoid of virus. These substances were prepared in a GMP-accredited facility, provided in prefilled syringes (3 μg in 0□5 mL), and administered intramuscularly on Days 1 and 14 (+14-day window) from participation onset.

Comprehensive assessments, encompassing medical history, physical examination, vital signs, and laboratory parameters, were conducted at baseline. Respiratory swabs for SARS-CoV-2 assessment were collected at screening and upon suspected COVID-19. Antibodies against SARS-CoV-2 were measured at various intervals. Participants were instructed to report the occurrence of symptoms suggestive of COVID-19, and were monitored post-vaccination. Follow-up visits were conducted electronically, by telephone, or in-person, during which adverse events and suspected COVID-19 were evaluated. The DSMB oversaw safety data.

A case of COVID-19 was defined, according to the U.S. Food and Drug Administration (FDA) criteria, as any of the following symptoms for at least 2 days, with a positive SARS-CoV-2 reverse transcription polymerase chain reaction (RT-PCR): fever or chills, cough, dyspnea, fatigue, muscle or body pain, headache, sore throat, nasal congestion or runny nose, nausea or vomiting, or diarrhea.^13^ Serum antibodies against the receptor binding domain (RBD) and viral nucleocapsid (N) were assessed using an electrochemiluminescence immunoassay (Elecsys, Roche) following the manufacturer’s instructions. Adverse events were classified and graded based on the guidance from the FDA^14^ and the U.S. National Cancer Institute criteria.^15^ All potential cases of COVID- 19 were to be followed until resolution. Investigators recorded symptom scores during follow-up according to the World Health Organization (WHO) Progression Score.^16^

### Outcomes

The primary efficacy endpoint was the incidence of RT-PCR-confirmed symptomatic COVID-19 cases commencing at least 14 days after the second dose of CoronaVac or placebo. Secondary efficacy endpoints encompassed symptomatic cases according to previous exposure to SARS-CoV-2, WHO Clinical Progression Scale, and severity of the disease. Prior SARS-CoV-2 exposure was defined as any positive/reactive test in one of three sources: a test of nucleic acid detection of SARS-CoV-2 in a clinical sample prior to or on the day of the first vaccination, or by positive test of antibodies against SARS-CoV-2 on the day of first vaccination. A subject was considered to not been previously exposed if tests were negative/non-reactive and both external PCR and IgT/RBD were available.

The primary safety endpoint was the frequency of solicited and unsolicited local and systemic adverse reactions with onset within 7 days from vaccination, according to the age groups (18-59 of age and ≥ 60 years). Adverse reactions were defined as adverse events considered to have a reasonable causal relationship (either possibly, probably, or definitely related) to vaccination. Secondary safety endpoints included: solicited and unsolicited adverse reactions after each dose with onset within 7 days from vaccination, solicited and unsolicited adverse reactions after each dose and after any dose with onset within 28 days from vaccination, frequency of cases of adverse events of special interest in participants who received at least one dose of the experimental product, and unsolicited adverse reactions of special interest (see Protocol for the list of events that were considered of special interest). Safety outcomes were analyzed in all participants (i.e., As-Treated population) and by age groups. A complete list of the study endpoints and their description is provided in the Protocol.

### Statistical analysis

The Statistical Analysis Plan (SAP) is provided in the Supplementary Material. The primary analysis was conducted in the Per-Protocol population, defined as all randomized participants who did not violate the protocol and received the two doses of CoronaVac or placebo. All cases were assessed by a blinded independent clinical endpoint adjudication committee up to December 16, 2020, the cutoff date for the primary analysis of efficacy used for EUA, and up to July 29, 2021, the cut-off data for the final analysis. Sample size computation for this group-sequential design^17^ was facilitated by the gsSurv function in the R package (R Foundation for Statistical Computing, Vienna, Austria, https://www.R-project.org/)^18^, gsDesign (gsDesign: Group Sequential Design_. R package version 3.5.0, https://CRAN.R-project.org/package=gsDesign)^19^ using the method of Lachin and Foulkes, with the Hwang-Shih-DeCani spending function and a single interim analysis when 40% of events were accrued.^20^ At least 151 primary endpoints were needed for the primary analysis, with the DSMB conducting an interim analysis after at least 61 events accrued.

A total of 13,060 participants were needed to demonstrate that the lower limit of the 95% confidence interval (CI) for vaccine efficacy (VE) was above 30%, assuming an attack rate of 2’115% over 12 months, power of 90%, one-sided type-I error of 2’115%, and a 5% annual dropout rate. VE was defined as 1 minus the hazard ratio (HR) from a Cox regression stratified by age group, with inference derived from Wald confidence limits and p-values. For the primary analysis of efficacy, participants were right-censored at the occurrence of a qualifying major protocol deviation, study drop-out not related to COVID-19, suspected but not laboratory-confirmed COVID-19, non-study COVID-19 vaccination, or the date of individual unmasking (in the case of the final analysis that followed the EUA). Only first events (per person) were considered as primary endpoints; repeat endpoints were not analyzed. The incidence of the primary endpoint was computed using the Kaplan-Meier method. The final efficacy analysis was foreseen after at least 80% of the participants were individually unmasked after the EUA.

The intention-to-treat (ITT) population comprised all participants who received at least one dose of CoronaVac or placebo. A modified ITT population comprised all participants who received both doses of CoronaVac or placebo, regardless of any protocol deviation. The safety population included all participants who received at least one dose of the vaccine or placebo and had safety data available. Solicited adverse events and reactions were analyzed in the respective reactogenicity population, defined as all participants in the safety population who received the dose of interest and returned the diary vaccine card. Solicited events included those observed by the clinical site within 60 minutes of injection and those recorded in the participants’ diaries. Secondary analyses included those related to safety, the incidence of the primary endpoint in the ITT population, and others described in the SAP provided in the Supplementary Material. Sensitivity analyses of efficacy were conducted based on selected baseline characteristics. Statistical analysis was conducted using SAS 9.4 and R 4.2.1 for the figures.

## RESULTS

Figure S1 in the Supplementary Material provides a chronological overview of key dates throughout the study period. For the interim analysis, the data cut-off was November 27^th^, 2020, and there were 120 virologically confirmed cases occurring >14 days from the second vaccination submitted for adjudication. Among these, there were 107 valid cases, but with two major protocol deviations leading to exclusion, yielding 105 for analysis, deriving a result of efficacy of 58.50% (95% CI, 29·88 to 75·44), 31 cases in the vaccine group (11·7 per 100 person-years; 95% CI, 11·55 to 24·13) and 74 cases (40·83 per 100 person-years; 95% CI, 32·06 to 51·26) in the placebo group. Based on these results, the DSMB suggested to continue with the trial as planned. For the primary efficacy analysis used to obtain EUA for CoronaVac in Brazil, data cut-off was December 16, 2020. During the period from July 21 to December 16, 2020, a total of 12,571 participants were screened, leading to randomization of 12,270 individuals (Figure S2). Demographic characteristics of the per-protocol set are outlined in Table S1. Among participants included in the primary analysis, a total of 252 confirmed symptomatic COVID-19 cases were identified. Of these, 85 cases in the vaccine group (11·7 per 100 person-years; 95% CI, 9·38 to 14·52) and 167 cases (23·6 per 100 person-years; 95% CI, 20·19 to 27·51) in the placebo group. This translated to a vaccine efficacy of 50·39% (95% CI, 35·26% to 61·98%; *p*=0·0049) in preventing symptomatic COVID-19 (Table 1 and Figure 2A). These findings led to the issuance of EUA for CoronaVac in January 2021.

**Table 1:** Vaccine efficacy of CoronaVac against COVID-19 2 weeks after the second dose vaccination for the primary and selected secondary efficacy endpoints in the primary analysis for Emergency Use Authorization (cutoff on December 16, 2020) and in the final analysis (cutoff on July 29, 2021) according to per-protocol population.

| Total (N= V/P) | Outcome | Vaccine Efficacy [95% CI] | Cases distribution<br>Vaccine/Placebo | *p* * |
| --- | --- | --- | --- | --- |
| **Cutoff on December 16, 2020 (N= 4653/4589)** |  |  |  |  |
| Primary endpoint | COVID-19 case | 50 □ 39% [35 □ 26%,<br>61 □ 98%] | Total 85/167, Elderly 2/3 | **0 □ 0049** |
| Secondary endpoints | WHO score ≥ 3 | 77 □ 96% [49 □ 94%, 90 □ 29%] | Total 7/31, Elderly 0/1 | **0 □ 0029** |
|  | WHO score ≥ 4 | 100 □ 00% [NC, 100 □ 00%] | Total 0/7, Elderly 0/1 | 0 □ 4967 |
|  | Severe case | 100 □ 00% [NC, 100 □ 00%] | Total 0/5, Elderly 0/0 | 0 □ 4972 |
| **Cutoff on July 29, 2021 (N= 5862/5758)** |  |  |  |  |
| Primary endpoint | COVID-19 case | 44 □ 58% [34 □ 89%,<br>52 □ 83%] | Total 233/405, Elderly 6/7 | **0 □ 0023** |
| Secondary endpoints | WHO score ≥ 3 | 82 □ 14% [64 □ 93%,<br>90 □ 90%] | Total 10/54, Elderly 0/3 | **<0 □ 0001** |
|  | WHO score ≥ 4 | 100 □ 00% [NC, 100 □ 00%] | Total 0/17, Elderly 0/3 | 0 □ 4949 |
|  | Severe case | 100 □ 00% [NC, 100 □ 00%] | Total 0/10, Elderly 0/2 | 0 □ 4961 |

CI, confidence interval; N, number of participants; P: participants in the placebo group; V: participants in the vaccine (CoronaVac) group; WHO: World Health Organization Clinical Progression Scale
\* p-value 'Pr < z' is the one-sided p-value to test against a HR of 0 □ 7 (VE of 30%).

The final analysis cutoff date for the PROFISCOV study was July 29, 2021, with more than 90% of participants unmasked. From July 21, 2020, to July 29, 2021, a total of 13,166 participants provided informed consent, of which 12,688 were randomized (6,344 in each group). Figure 1 shows the study flowchart for the final data cutoff. Baseline characteristics of randomized participants who had received at least one dose of vaccine or placebo are summarized in Table 2. Participants had a mean age of 39·6 years, 93% were 18-59 years, 63·7% were female, and 75·6% were self-reported as white. Previous exposure to SARS-CoV-2 was demonstrated in 9·5%. Baseline features of the per-protocol population at the time of the final analysis are displayed in Table S2. A total of 807 cases of COVID-19 were identified. Among the per-protocol population, 638 confirmed COVID-19 cases occurred, with 233 cases in the vaccine group (17·06 per 100 person-years; 95% CI, 14·94 to 19·39) and 405 in the placebo group (30·78 per 100 person-years; 95% CI, 27·86 to 33·93), as shown in Table 1. This translated to a vaccine efficacy of 44.58% [95% CI, 34·89% to 52·83%; *p*= 0·0023]. The efficacy of CoronaVac in preventing COVID-19 severity (score ≥3) was 82·14% (95% CI, 64·93% to 90·90%; *p*<0·0001; Table 1 and Figure S3). Vaccine efficacy was observed to be lower among elderly (16.3% CI, -149.0% to 71.9%) than younger participants (45.1%, CI 35.4% to 53.3%), although few cases were available for analysis among the elderly.

**Figure 1:**
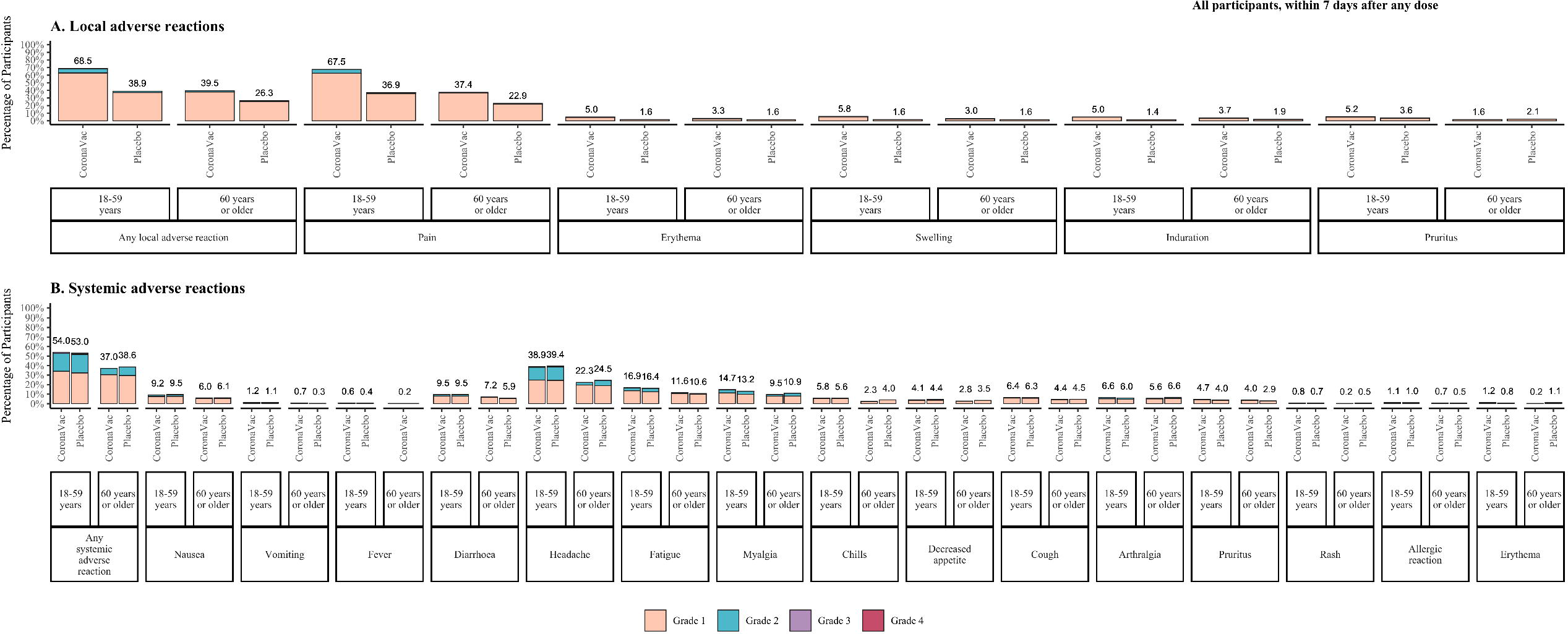
CONSORT flowchart of the study population for the final analysis at data cutoff July 29, 2021

**Figure 2:**
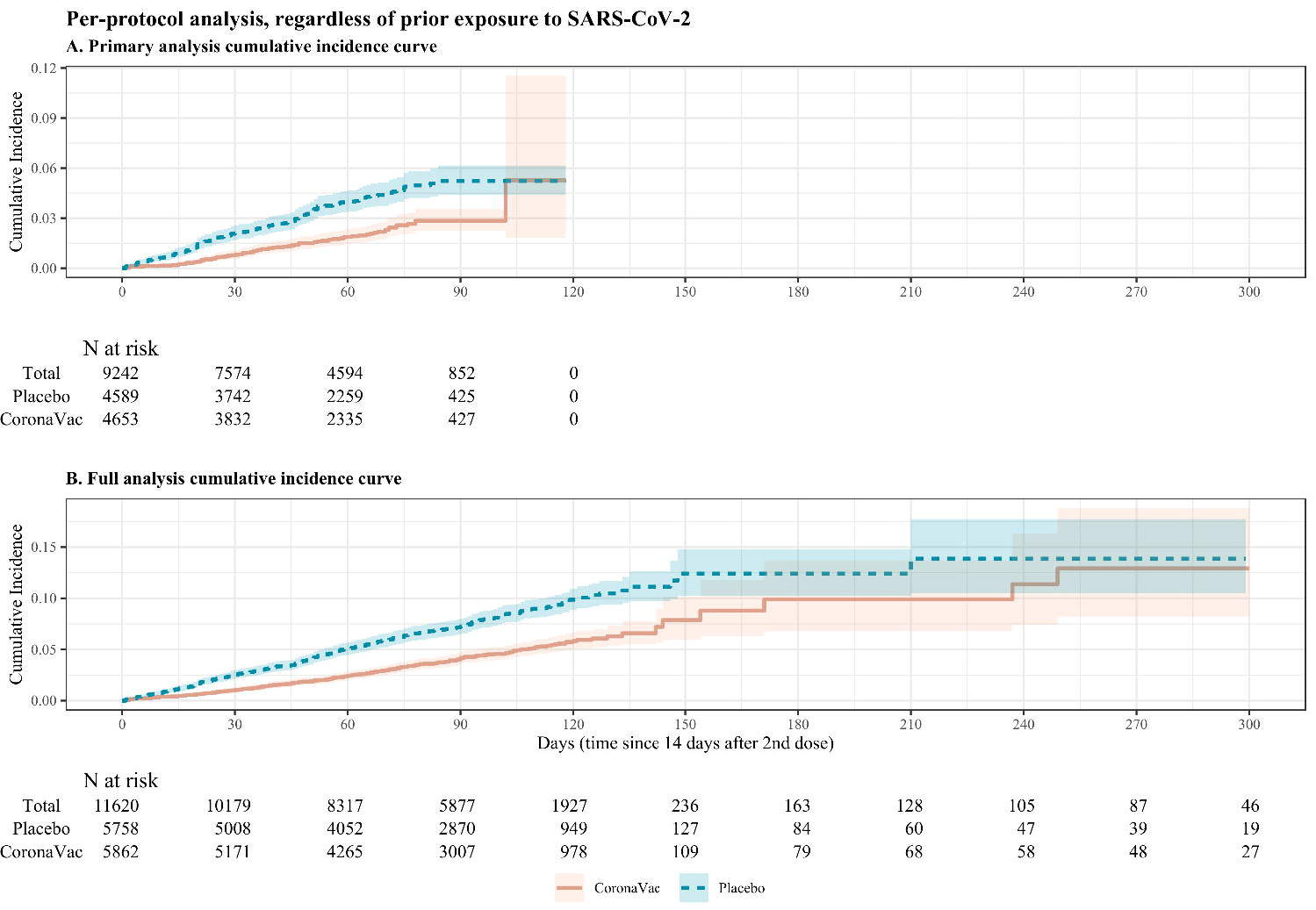
Cumulative incidence of symptomatic COVID-19 according to per-protocol population regardless of age and prior exposure to SARS-CoV-2. Caption: Panel A: Cumulative incidence of RT-PCR confirmed symptomatic COVID- 19 cases with onset at least 14 days after the second dose of CoronaVac or placebo in the per-protocol population, as of the data cutoff December 16, 2020. Panel B: Cumulative incidence of RT-PCR confirmed symptomatic COVID-19 cases with onset at least 14 days after the second dose of CoronaVac or placebo in the per-protocol population, as of the data cutoff July 29, 2021. Vaccine efficacy estimated as 1-HR through Cox regression model stratified by age group for time to COVID-19 onset and 95% confidence intervals were estimated using Wald method.

**Table 2:**
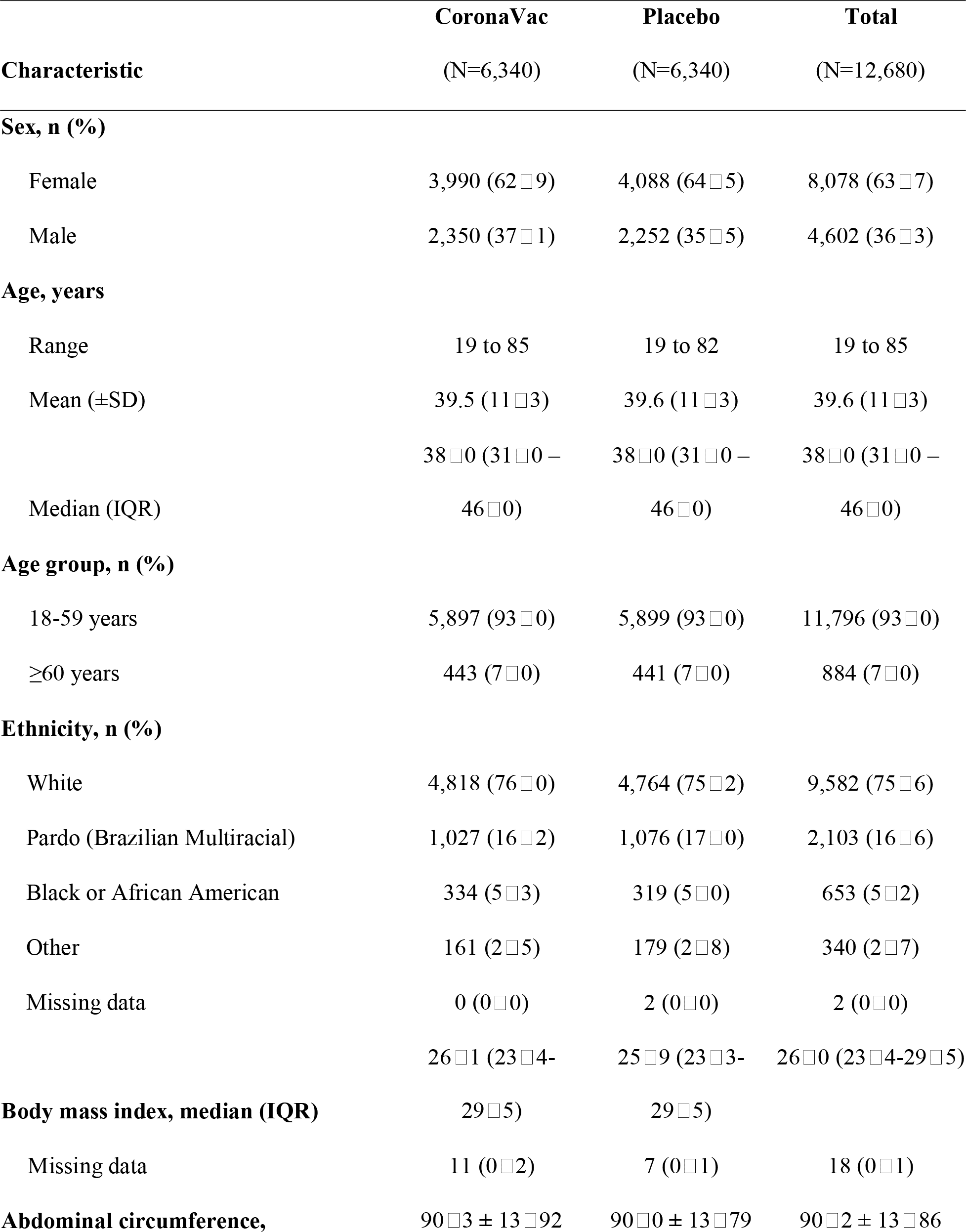
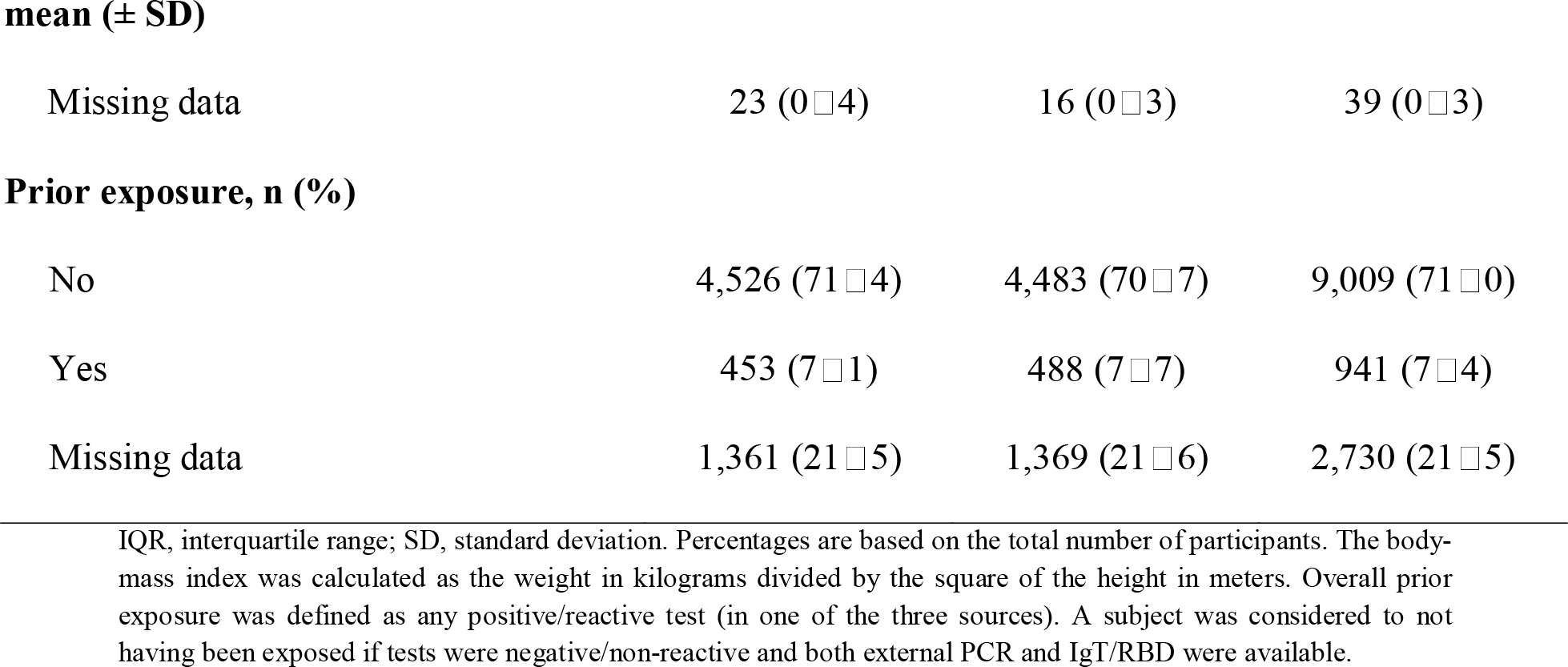
Baseline socio-demographic characteristics of the randomized participants who received at least one dose of CoronaVac or placebo (N=12,680)

|  | CoronaVac | Placebo | Total |
| --- | --- | --- | --- |
| Characteristic | (N=6,340) | (N=6,340) | (N=12,680) |
| **Sex, n (%)** |  |  |  |
| Female | 3,990 (62□9) | 4,088 (64□5) | 8,078 (63□7) |
| Male | 2,350 (37□1) | 2,252 (35□5) | 4,602 (36□3) |
| **Age, years** |  |  |  |
| Range | 19 to 85 | 19 to 82 | 19 to 85 |
| Mean (±SD) | 39.5 (11□3) | 39.6 (11□3) | 39.6 (11□3) |
|  | 38□0 (31□0 – | 38□0 (31□0 – | 38□0 (31□0 – |
| Median (IQR) | 46□0) | 46□0) | 46□0) |
| **Age group, n (%)** |  |  |  |
| 18-59 years | 5,897 (93□0) | 5,899 (93□0) | 11,796 (93□0) |
| ≥60 years | 443 (7□0) | 441 (7□0) | 884 (7□0) |
| **Ethnicity, n (%)** |  |  |  |
| White | 4,818 (76□0) | 4,764 (75□2) | 9,582 (75□6) |
| Pardo (Brazilian Multiracial) | 1,027 (16□2) | 1,076 (17□0) | 2,103 (16□6) |
| Black or African American | 334 (5□3) | 319 (5□0) | 653 (5□2) |
| Other | 161 (2□5) | 179 (2□8) | 340 (2□7) |
| Missing data | 0 (0□0) | 2 (0□0) | 2 (0□0) |
|  | 26□1 (23□4- | 25□9 (23□3- | 26□0 (23□4-29□5) |
| **Body mass index, median (IQR)** | 29□5) | 29□5) |  |
| Missing data | 11 (0□2) | 7 (0□1) | 18 (0□1) |
| **Abdominal circumference,** | 90□3 ± 13□92 | 90□0 ± 13□79 | 90□2 ± 13□86 |
| **mean ( ± SD)** |  |  |  |
| Missing data | 23 (0□4) | 16 (0□3) | 39 (0□3) |
| **Prior exposure, n (%)** |  |  |  |
| No | 4,526 (71□4) | 4,483 (70□7) | 9,009 (71□0) |
| Yes | 453 (7□1) | 488 (7□7) | 941 (7□4) |
| Missing data | 1,361 (21□5) | 1,369 (21□6) | 2,730 (21□5) |

IQR, interquartile range; SD, standard deviation. Percentages are based on the total number of participants. The body-mass index was calculated as the weight in kilograms divided by the square of the height in meters. Overall prior exposure was defined as any positive/reactive test (in one of the three sources). A subject was considered to not having been exposed if tests were negative/non-reactive and both external PCR and IgT/RBD were available.

Vaccine efficacy was similar in the ITT population (Tables S3 and S4, and Figure S4). Subgroup analysis of vaccine efficacy in the per-protocol and intention-to-treat populations are summarized in Figure S5.

Regarding the safety analyses, all 12,680 participants were monitored for adverse events from the time of vaccination until the date of dropout or data cut off for analysis. An overview of the adverse events and reactions after any dose is shown in Table S5. Solicited adverse events were reported more frequently in the CoronaVac group than in the placebo group. Most events were mild to moderate in severity. Adverse events were frequent in both groups, and more frequent in the CoronaVac than in the placebo group within 7 days after any dose (81□6% *vs*. 71□5%). Similar results were observed within groups of participants aged 18-59 and those aged ≥60 years.

Solicited local and systemic adverse reactions within 7 days after any dose were more common in the vaccine group, with pain at injection site and headache being the most frequently reported reactions (Figure 3). Solicited adverse reactions within 7 days of any dose were more frequently reported in the vaccine group than in placebo in the overall population (78□4% *vs*. 65□4%; *p*<0□0001) and in participants aged 18-59 years (80□1% *vs*. 66□5%, *p*<0□0001), and at similar rates for participants aged ≥60 (55□1% *vs*. 48□7%; *p*=0□0772). Overall, solicited local and systemic adverse reactions were less common in older participants (aged ≥60 years). Adverse reactions reported within 7 days after each dose are provided in Figure S6 and S7.

Reactions in the CoronaVac group were Grade 1 in severity in most cases. Pain was the most common local reaction in both groups, being reported with a significantly higher frequency among CoronaVac recipients than in those in the placebo group within 7 days of any dose (65□4% and 36□1%), of dose 1 (47□5% vs 24□4%) and of dose 2 (49□8% vs 21□2%), respectively.

Solicited systemic adverse reactions within 7 days after any dose occurred at similar frequencies among participants in the vaccine and placebo groups in the overall population (52□8% *vs*. 52□1%) and in participants aged 18-59 years (54□0% *vs*. 53□0%) and ≥60 years (37□0% *vs*. 38□6%). In both groups, most common systemic reactions were headache, fatigue, myalgia, nausea, and diarrhea. Regarding the occurrence of systemic solicited adverse reactions within 7 days of the first dose, the frequency of arthralgia, chills, cough, decreased appetite, diarrhea, fatigue, fever, headache, hypersensitivity, myalgia, nausea, rash, systemic erythema, systemic pruritus, and vomiting did not significantly differ between recipients of CoronaVac and those who received placebo.

Among the secondary safety endpoints, the frequency of adverse reactions to the vaccine, solicited and unsolicited, after each of the two doses within the period of 4 weeks after vaccination, according to the age group, adults (18-59 years old), and elderly (60 years or older) subjects was also assessed (Figures S8, S9 and S10). Rates were somewhat higher for the longer observation period, but otherwise similar to results reported within 7 days.

Ten cases of confirmed severe COVID-19 were reported in placebo recipients, while no case of confirmed severe COVID-19 occurred among vaccine recipients. Nineteen pregnancies occurred within the safety follow-up period (11 in the vaccine and 8 in the placebo group). A total of 5 abortions were reported (2 in the vaccine and 3 in the placebo group). None of them were considered related to the vaccine.

Adverse events of special interest were reported for 2□7% and 4□6% of participants in vaccine and placebo groups, respectively. A summary of adverse events of special interest overall and by System Organ Class (SOC) and Preferred Term is shown in Table S10. Nervous systems disorders was the SOC with the largest number of events reported, and within this category anosmia (reported in 1□7% in the vaccine group and 2□9% in placebo group) and ageusia (1□1% in the vaccine group and 1□7% in placebo group) were the most common Preferred Terms in both groups.

Serious adverse events (SAE; Table S9) after any dose were reported by 110 (0□9%) participants, 48 (0□8%) in the vaccine group and 62 (1□0%) in the placebo group (p=0□2130). Only one SAE (ulcerative colitis), reported between 3-days after vaccination, was considered as possibly related to the investigational vaccine.

Four deaths were reported in this study (0□03% of participants). One fatal adverse event occurred in the vaccine group (suicide), and three in the placebo group (COVID- 19, severe acute respiratory syndrome, and cardiopulmonary arrest). None of them were related to the investigational vaccine. No case of vaccine-associated worsened respiratory disease was reported in the vaccine group.

## DISCUSSION

This study sought to evaluate the safety and efficacy of CoronaVac among HCWs in Brazil, a population with elevated exposure risk due to their direct contact with COVID- 19 patients. Our findings indicate that the vaccine provided an overall protection rate of 50.39% against symptomatic COVID-19 at the primary cutoff date. Subsequent analysis, encompassing data up to July 29, 2021, showed vaccine efficacies of 44□58%, 82□14%, 100%, and 100% against symptomatic COVID-19, WHO scores of ≥3 and ≥4, and severe disease, respectively. In both evaluations, CoronaVac met the *a priori* criteria for vaccine efficacy, which mandated that the lower limit of the 95% CI exceeds 30%. The vaccine demonstrated favorable tolerability without any alarming safety indications.

Earlier phase 3 trials yielded varying efficacy estimates for CoronaVac in preventing PCR-confirmed symptomatic COVID-19, 14 days after the second dose, with rates of 65□3% in Indonesia and 83□5% in Turkey.^7,8^ The discrepancies in efficacy may be attributed to differences in study designs, populations, and statistical methodologies between these trials and ours. Interestingly, in a pos-hoc analyze, we observed that vaccine efficacy among those that received the two doses more than 21 days was higher than those with interval between doses less than 21 days (Fig S5). Real-world vaccine effectiveness echoed the outcomes from controlled studies, underscoring the vaccine’s demonstrated ability in preventing symptomatic disease, hospital admissions, ICU admissions, and COVID-19-related fatalities.^21–23^

Globally, over 40 countries and territories have granted authorization to CoronaVac, with its inclusion in the WHO Emergency Use Listings (EUL).^24^ Drawing from evidence available in June 2021, WHO advocated for the administration of CoronaVac in adults aged 18 and above, following a two-dose regimen spaced between 2 to 4 weeks. The absence of an upper age limit for the vaccine was justified by compelling immunogenicity and safety data, suggesting comparable protection and safety profiles in older and younger demographics. In November 2022, a decision to extend the age indication, allowing primary immunization for individuals aged 3-17 years, was reached.^25^ This decision was based on WHO’s data review and the endorsement from the National Medical Products Administration, the official WHO regulatory authority for this vaccine.

During the nascent stages of the COVID-19 pandemic, the WHO recommended a minimum vaccine efficacy threshold of 50% for a vaccine to be deemed efficacious. CoronaVaćs definitive market authorization was challenged by the predefinition of this threshold, especially when referencing the final data (vaccine efficacy against RT-PCR-confirmed symptomatic disease: 44□58% [34□89%, 52□83%]). Every vaccine approval process inherently entails a balancing act between benefits and potential risks. In certain scenarios, even limited protection can be deemed acceptable, particularly if the illness in question is severe and lacks alternative protective measures. Robust data from controlled and real-world effectiveness studies reinforce CoronaVac’s efficacy in thwarting severe diseases, hospitalizations, and fatalities.^26^ While safeguarding against symptomatic disease remains a paramount objective, a vaccine’s ability to mitigate severe disease, akin to influenza vaccines, represents a transformative impact on public health. With the emergence of variants like omicron and its descendants, which potentially bypass immunity, a vaccine’s capability to protect against severe illness becomes an even more pivotal feature in pandemic control.

Our study has limitations. The primary emphasis on Brazilian HCWs could constrain the generalizability of our findings to broader populations. During the study duration the main variants that circulated were the Wuhan beta and Delta strains, all of which might not be relevant nowadays due to the predominance of omicron-related sub-lineages. It’s also worth noting that prior exposure to SARS-CoV-2 was confirmed in only 9□5% of our participants, which may not represent wider population exposure rates accurately.

In conclusion, CoronaVac has emerged as a promising option for COVID-19 vaccination option, exhibiting a commendable safety profile and moderate protective efficacy against symptomatic manifestations of the disease in Brazilian healthcare workers. Although adverse events were slightly higher in the vaccinated group, their predominantly mild nature underscores the vaccine’s safety. The vaccine demonstrated significant potential for pandemic control due to its easy of deployment and lower cost compared to other emerging vaccine platforms available early in the pandemic, which was desperately needed globally. Further research is warranted to elucidate the impact on immunogenicity and safety profile of XBB-updated versions of the vaccine used as a booster vaccination.

## Contributors

JM wrote the first draft of the paper; EP, PB, PP, CG participated in the statistical analysis, wrote the Statistical Analysis Plan, and drafted the figures and tables, RP contributed substantially to the drafting of the study protocol and the primary analysis of vaccine efficacy, conducted with a data cutoff on December 16, 2020; all the authors contributed substantially to data collection and reviewed the draft; all authors read and approved the final version of the manuscript.

## Data sharing

Individual de-identified data on trial participants, and the data dictionary will be made available to qualified investigators following a request for use of these materials, and will be held at the Instituto Butantan. Request for access to trial data should be addressed to the corresponding author.

## Acknowledgements

The authors would like to thank the participants who enrolled in the study. The authors wish to also thank the investigators, study site staff, CRO team (IQVIA Brazil and Assign Data Management and Biostatistics - AssignDMB -, Innsbruck, Austria), PATH staff and the sponsor personnel for their contributions to the implementation and conduction of the trial. Special thanks to former and current Instituto Butantan employees who were crucial to make this trial happen, and to João Italo Dias França, M.Sc., for his contribution reviewing the final results. The trial was conducted by Instituto Butantan, São Paulo, Brazil. Ana Elisa B. Bueno da Silva, PhD (DENDRIX, Sao Paulo, Brazil), provided medical writing support for this publication, which was funded by Fundacao Butantan.

---

Figure 3: Frequency of solicited local and systemic adverse reaction within 7 days of any dose by age group

